# Identification of causal genes and mechanisms by which genetic variation mediates juvenile idiopathic arthritis susceptibility using functional genomics and CRISPR-Cas9

**DOI:** 10.1101/2025.05.22.25325739

**Authors:** Antonios Frantzeskos, Valeriya Malysheva, Chenfu Shi, Danyun Zhao, Muskan Gupta, Stefano Rossi, James Ding, CLUSTER consortium, Wendy Thomson, Steve Eyre, John Bowes, Mikhail Spivakov, Gisela Orozco

## Abstract

**Objective:** Genome-wide association studies (GWAS) have identified numerous single nucleotide polymorphisms (SNPs) associated with juvenile idiopathic arthritis (JIA), the majority of which are located in non-coding regions such as enhancers. This presents a challenge for pinpointing causal variants and their target genes. Interpreting these loci requires functional genomics data from disease-relevant tissues, which has been lacking for JIA. This study seeks to fill that gap and elucidate the biological mechanisms underlying JIA susceptibility.

**Methods:** We performed low-input whole genome promoter Capture Hi-C (PCHi-C) and ATAC-seq on CD4+ T cells from three JIA oligoarthritis patients. To link JIA-associated SNPs to potential causal genes, we integrated PCHi-C data with JIA GWAS summary statistics using our Bayesian prioritisation algorithm, Capture Hi-C Omnibus Gene Score (COGS). ATAC-seq was used to further annotate JIA GWAS loci in CD4+ T cells. We then employed CRISPR activation and interference (CRISPRa/i) in Jurkat cells to validate the prioritised SNPs and their corresponding genes.

**Results:** Chromatin interactions between JIA-associated SNPs and gene promoters were identified in 19 of 44 non-MHC JIA loci, linking 61 known and novel target genes to the disease. Through COGS, we prioritised seven putative causal genes for JIA: *RGS14, ERAP2, HIPK1, CCR4, CCRL2, CCR2*, and *CCR3*. SNPs within promoter-interacting regions (PIRs) of these genes were further validated using CRISPRa/i to confirm their roles in regulating gene expression.

**Conclusions:** This study provides insights into the genetic architecture of JIA by integrating genomic and epigenomic data, identifying disease-related genes, functionally validating risk SNPs, and highlighting candidate drugs for repurposing.

**Key messages:** *What is already known on this topic:* Recent genome-wide association studies in JIA have identified genetic loci associated with disease risk. However, the precise mechanisms by which these variants contribute to disease pathology remain unclear, as most do not directly alter protein-coding genes. It has been proposed that non-coding SNPs can affect genes that are important in disease through disruption of enhancer-mediated regulatory mechanisms that control their expression, with enhancers exerting their effects through chromatin interactions. Functional characterisation of risk loci is essential to delineate causal SNPs and target genes in JIA.

*What this study adds:* This study is the first to utilise low-input Promoter Capture Hi-C to map long-range chromatin interactions in CD4+ T cells from JIA patients, alongside ATAC-seq to assess chromatin accessibility within the same samples. It identifies 61 potential target genes at JIA-associated loci and validates the regulatory roles of some of these through CRISPR activation and interference. This work enhances our understanding of how genetic variants modulate gene expression in immune cells, shedding light on key pathways involved in JIA pathogenesis.

*How this study might affect research, practice or policy:* Highlights new potential causal genes in JIA which can help understand the pathological mechanisms in JIA, and suggests the potential to repurpose CCR2/CCR5 inhibitors in JIA.

## Introduction

Juvenile idiopathic arthritis (JIA) is the most common chronic paediatric rheumatic disease, characterised by unexplained joint inflammation. Despite extensive research, its pathogenesis remains largely elusive. Similar to other complex diseases, JIA is speculated to arise from an aberrant immune response in genetically susceptible individuals [1]. Understanding its genetic basis is crucial, as it can facilitate improved diagnosis, inform personalised treatment strategies, and aid in the identification of novel or repurposed therapeutic targets.

Genome-wide association studies (GWAS) have expanded our understanding of JIA genetics, with the largest recent GWAS identifying 44 susceptibility loci [2]. However, as with many complex diseases, lead GWAS SNPs in JIA are typically in linkage disequilibrium (LD) with other variants and predominantly reside in non-coding regions. These non-coding SNPs often localise in DNA regulatory elements, and may impact normal gene expression in a cell-type- and stimulus-specific manner [3–5]. Notably, JIA susceptibility SNPs are enriched in enhancers active in various T cell subsets, indicating their functional relevance in disease pathogenesis [2]. To translate GWAS findings into tangible patient benefits, it is essential to link risk SNPs to their target genes and understand their influence on gene regulation.

A key challenge in JIA research has been the limited availability of disease-relevant functional datasets, largely due to the difficulty of obtaining biological material from paediatric patients. To address this, we employed low-input Promoter Capture Hi-C (PCHi-C) to generate the first chromatin interaction maps from CD4+ T cells of JIA patients, enabling the interrogation of regulatory interactions between non-coding SNPs and gene promoters. We integrated this with ATAC-seq from the same samples to assess chromatin accessibility and applied our Bayesian Capture Hi-C Omnibus Gene Score (COGS) approach to prioritise candidate causal SNPs and genes.

To validate the functional relevance of selected SNPs and demonstrate SNP-gene relationships we utilised CRISPR activation and interference (CRISPRa/i) in the Jurkat T-lymphocyte cell line. This study provides the first functional validation of JIA-associated SNPs using CRISPRa/i and establishes an initial mechanistic link between JIA risk variants and causal genes. Among the regulatory interactions identified, SNPs at the *CCR3* locus illustrate how non-coding variants modulate gene expression, highlighting a locus with potential for drug repurposing [6,7]. These findings contribute to bridging the gap between genetic associations and functional mechanisms in JIA, offering insights that could inform future therapeutic strategies.

## Methods

### Patient Recruitment and CD4+ T cell isolation

Participants, were recruited under ethics approval granted by the NHS Health Research Authority North West-Greater Manchester West Research Ethics Committee (REC reference: 99/8/084, IRAS ID: 32231). The Committee approved the study, and all participants consented. Peripheral blood mononuclear cells (PBMCs) were isolated using Ficoll density gradient centrifugation. 15ml of Ficoll (Sigma Aldrich) was added per Leucosep tube (Thermo Scientific) and centrifuged at 1000xg for 30sec to allow the Ficoll to move below the filter. Subsequently 30-35ml of blood was added to each Leucosep tube and centrifuged at 1200xg for 30min without any deceleration to avoid disruption of the PBMC layer. The top layer of plasma was removed and the PBMC containing layer was then collected in a new 50ml falcon tube and washed using phosphate buffered saline (PBS) (Thermo Scientific), 2% fetal bovine serum (FBS) (Thermo Scientific), 2mM (EDTA) (Thermo Scientific) and centrifuging at 500xg for 10min. This was repeated until the supernatant was clear. Subsequently EasySep Human CD4^+^ T cell negative selection kits (StemCell #17952) were used to isolate CD4^+^ T cells.

### Low input Promoter Capture Hi-C

We combined PCHi-C [8] with Hi-C library generation as described previously [9], with some modifications. Per replicate, 250,000 cells were fixed in 2% PFA for 10 minutes, lysed in lysis buffer (30 minutes on ice), and digested with DpnII (NEB) overnight at 37°C rotating (950rpm). Restriction overhangs were filled-in with Klenow (NEB) using biotin-14-dATP (Jena Bioscience), and ligation was performed in ligation buffer for 4 hours at 16°C (T4 DNA ligase; Life Technologies). After overnight decrosslinking at 65°C, the ligated DNA was tagmented to produce fragments of 300-700 bp range. Ligation products were isolated using MyOne C1 streptavidin beads (Life Technologies), followed by washing with Wash&Binding buffer and nuclease-free water. Isolated Hi-C ligation products on the beads were then used directly for PCR amplification, and the final Hi-C library was purified with AMPure XP beads (Beckman Coulter). Promoter Capture Hi-C was performed using a custom-design Agilent SureSelect system following the manufacturer’s protocol.

### PCHi-C data pre-processing and detection of significant interactions

Sequencing data from three PCHi-C biological replicates were aligned to the hg38 genome assembly using Bowtie2 [10] and quality-controlled using HiCUP [11]. Significant interactions were then detected jointly across the replicates by CHiCAGO [12] as previously described [13] at single DpnII fragment resolution and in bins of fragments approximately 5kb in length, with the baited promoter fragments left solitary (unbinned). For CHiCAGO analysis at single-fragment resolution, p-value weights were estimated following our previously described procedure [13]; default p-value weights were used for the 5kb analysis. A CHiCAGO score cutoff of ≥5 was used for both resolutions to identify significant interactions.

### Putative causal gene prioritisation: COGS

To run COGS, we adapted the code from the R package rCOGS (https://github.com/ollyburren/rCOGS [github.com]) to use the data.table framework instead of GenomicRanges for optimised speed. We used GWAS summary statistics for JIA from López-Isac *et al*., study (https://www.ebi.ac.uk/gwas/studies/GCST90010715 [ebi.ac.uk]). We used linkage disequilibrium (LD) blocks calculated for GRCh38 from https://github.com/jmacdon/LDblocks_GRCh38128 [github.com] and minor allele frequencies from the 1000 Genomes Project, European individuals. Protein coding SNPs were identified using VEP version 99.2 (https://github.com/Ensembl/ensembl-vep [github.com]). We obtained gene transcription start sites (Havana and Ensembl/Havana merge) from Ensembl GRCh38 release 88 (March 2017), matching the version used to design the DpnII promoter capture system. We included promoters whether or not they were targeted in the capture system, enabling COGS to prioritise all gene targets where the causal variants were located near the gene promoter (defined as +/-5 DpnII fragments from the transcription start site) or within coding sequence. Promoter-interacting regions with CHiCAGO interaction scores ≥5 were used as COGS input. The results for each protein-coding gene were linked across datasets using Ensembl gene IDs as primary identifiers. The Major Histocompatibility Complex was removed (GRCh38 6:28510120-33480577), prior to running COGS.

### ATAC-seq

ATAC-seq libraries were prepared from 50,000 CD4+ T cells per JIA patient using the Omni-ATAC protocol as released by the Kaersten lab [14,15]. Library size was checked by TapeStation 4200 (Agilent) and quality control was done by Quantstudio (Life technologies) using the NEBNext® Library Quant Kit for Illumina (E7630). Sequencing was conducted on a NextSeq2000 platform, generating 50 bp paired-end reads at a target depth of 50 million reads per library.

ATAC-seq reads were quality filtered and trimmed and adapters were removed using fastp v0.20.1 [16]. Reads were then mapped using bowtie2 v2.4.4 [10] using the very-sensitive pre-set. Duplicates were removed using the MarkDuplicates functionality of picard tools (“Picard Toolkit,” 2019) and mitochondrial reads were removed, as well as removing low quality alignments (MAPQ).

### Functional annotation of JIA associated SNPs

The regulatory roles of individual SNPs identified in the most recent JIA GWAS [2] were investigated through annotation that incorporated publicly available ChIP-seq data from the Roadmap Epigenomics Consortium [17], EpiMap predicted immune cell related enhancers [18], as well as in-house generated ATAC-seq peaks and PCHi-C data from the 3 JIA CD4+ T cell samples. Moreover, we utilised the enhancer-gene links predicted by the EpiMap project from transcriptional correlation with epigenomic marks for all immune related enhancers [18]. Additionally, all SNPs were investigated for potential eQTLs in whole blood using the eQTLGen Consortium data found at https://www.eqtlgen.org/ [19]. JIA loci and SNPs were visualised using the WashU genome browser (https://epigenomegateway.wustl.edu).

### Cell culture

The T cell like Jurkat E6.1 leukaemic T-lymphoblast expressing dCas9-KRAB or dCas9-VP64 [20] were obtained from University of California Berkeley cell culture facility. Cells were cultured in RPMI-1640 medium supplemented with 2mM L-glutamine, 10% FBS, and 1% penicillin-streptomycin (P/S) (Sigma Aldrich) at 37°C, 5% CO₂, and maintained at 2 × 10⁵–8 × 10⁵ cells/mL. LentiX HEK293T cells (Clontech) were cultured in DMEM (high glucose) supplemented with 10% FBS and 1% P/S at 37°C, 5% CO₂ and maintained at 60–80% confluency.

### gRNA design

gRNAs were designed using the online tool CRISPOR (http://www.crispor.tefor.net) and selected based on highest predicted efficiency and specificity scores in addition to lowest predicted off target sites. A total of 3 gRNAs were selected for each SNP region targeted within 200 bp from the target SNP. For each gRNA forward and reverse oligos were ordered with *Esp3I* overhangs for cloning into the pLKO5.sgRNA.NeoR plasmid (adapted from the pLKO5.sgRNA.EFS.GFP (Addgene 57822)).

### Lentiviral production

Lentivirus was produced in LentiX HEK293T cells using a third-generation system comprised of pMD2.G (Addgene #12259), pMDLg/pRRE (Addgene #12251), and pRSV-REV (Addgene #12253). The transfer/gRNA plasmids were pooled, 2μg each (total 6μg) in addition to packaging plasmids; 1.5μg pMD2.G, 1.5μg pMDLg/pRRE and 3μg pRSV-REV. This was combined with 72μg Polyethylenimine (PEI). All mixes were done in triplicate. The PEI:plasmid mix was added dropwise to LentiX HEK293T and cultured for 72h at which point the supernatant containing the lentivirus was centrifuged at 1200rpm for 5 minutes at 4°C and filtered through a 45μm cellulose acetate vacuum (Corning).

### Lentiviral transduction of Jurkats

Jurkat-dCas9KRAB and VP64 cells were seeded at 2×10^6^ in 2ml complete RPMI-1640 with 8 μg/mL polybrene and transduced with 1ml of harvested lentivirus. 24h post addition of lentivirus, cells were cultured in selection media containing 900μg/ml Neomycin (Gibco) until control cells died out and then in maintenance media (450μg/ml Neomycin) for another 72h.

### RNA extraction

Cells were pelleted and processed using the RNeasy mini RNA extraction kit (Qiagen) according to the manufacturer’s instructions. Concentration was measured using the NanoDrop 2000 spectrophotometer (Thermo Scientific).

### Taqman real-time qPCR (RT-qPCR) and data analysis

Gene expression was quantified using TaqMan real-time PCR (Applied Biosystems) with the RNA-to-Ct 1-step kit. Reactions were performed in triplicate on the QuantStudio Flex 12K (Applied Biosystems). All Taqman gene expression assays are shown in **supplementary table 1.** Expression levels were normalised to **UBC**. Fold change in gene expression was subsequently calculated using the 2^−ΔΔCT^ method [21]. Statistical significance was assessed using unpaired t-tests.

## Results

### Promoter Capture Hi-C identifies gene targets at JIA risk loci

We carried out for the first time a low-input genome-wide PCHi-C in primary CD4+ T cells from 3 JIA oligoarthritis patients. Following our recently published guidelines, we performed interaction calling at *DpnII* fragment-level and 5kb resolution in order to cover a broad distance range of interactions [13]. Using the CHiCAGO pipeline [12], we identified 37728 and 72069 unique intrachromosomal significant interactions (CHiCAGO score ≥5), involving nearly 10,000 genes, at fragment and 5kb (with solitary baits) resolutions. The PIRs overlapped with risk SNPs (LD R^2^>0.8 to lead SNP) at 19 out of the 44 non-MHC JIA associated loci. Within these 19 loci, JIA SNPs interacted with 61 genes. Of these, 15 overlapped with 22 notable genes from López-Isac *et al*., defined based on proximity to the lead SNP. In contrast, PCHi-C uniquely identified 46 additional targets through promoter interactions. Newly identified genes are highlighted in bold in Table 1.

**Table 1.**
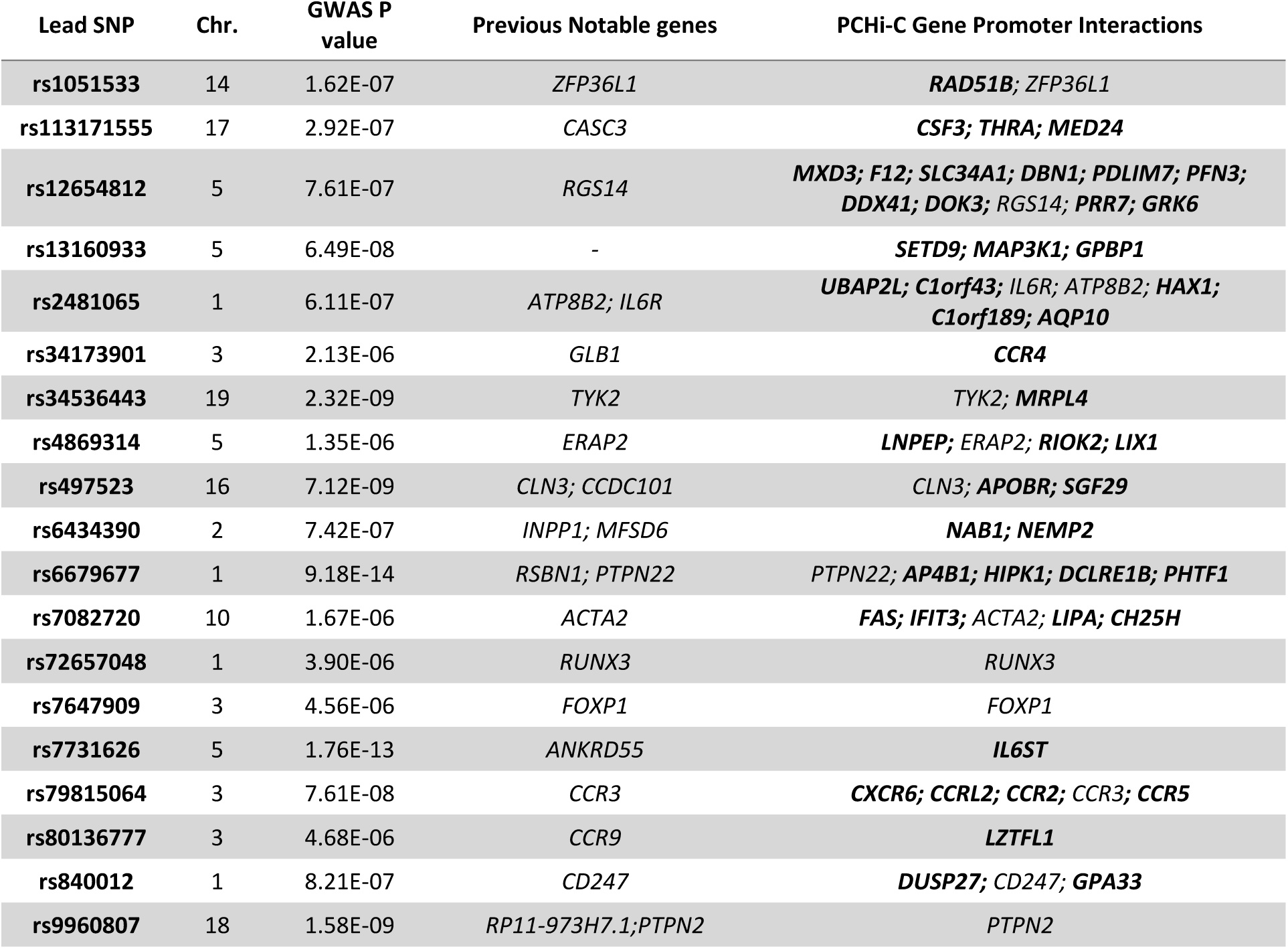
Genes identified by PCHi-C in primary CD4+ T cells from JIA patients that interact with JIA risk SNPs (LD R2>0.8 to lead SNP). Previously notable genes at these loci, as reported by López-Isac *et al*., are also shown.

### COGS prioritises PCHi-C target genes

We used the Bayesian prioritisation algorithm COGS [22] to prioritise putative causal genes in JIA for functional validation based on the identified promoter-SNP interactions. Briefly, COGS applies Bayesian statistical fine-mapping of GWAS SNPs that either interact with promoter regions of genes, reside in the promoter or in gene coding regions. We ran COGS on the latest JIA GWAS [2] and the PCHi-C interactions detected in this study at both fragment and 5kb resolution. This resulted in 7 genes within 5 loci above the posterior probability cutoff of 0.5 at 5kb resolution, shown in **Table 2**. Notably, several of the identified genes encode chemokine receptors, including *CCR4*, *CCRL2*, *CCR2*, and *CCR3*, which direct immune cells to inflamed tissues and have been implicated in autoimmune diseases, including JIA, by mediating leukocyte infiltration into synovial joints and sustaining inflammation [23–25]. Beyond chemokine receptors, PCHi-C identified additional genes of interest, including *ERAP2*, *HIPK1*, and *RGS14*. *ERAP2* plays a crucial role in antigen processing by trimming peptides for MHC class I presentation and has been associated with multiple autoimmune diseases [26]. A recent study demonstrated that rs2248374, an intronic SNP previously linked to JIA [27], regulates *ERAP2* expression through nonsense-mediated decay. Additionally, the study highlights the presence of independent regulatory signals in the region that may influence *ERAP2* expression. *HIPK1* is located within the *PTPN22* locus and was recently found to have lower expression in JIA CD4+ T cells compared to healthy controls, suggesting a possible role in immune dysregulation [28]. *RGS14* is a regulator of G-protein signalling, has primarily been studied in neuronal function [29,30] and warrants further investigation into its potential involvement in JIA pathogenesis.

**Table 2.** Top ranking (>0.5) COGS genes based on JIA CD4+ PCHi-C and risk SNPs from the López-Isac et al., GWAS.

| Ranking | Chr. | JIA Lead SNP | GWAS assigned gene (Based on proximity) | COGS prioritised Gene | COGS score |
| --- | --- | --- | --- | --- | --- |
| 1 | 5 | rs12654812 | RGS14 | RGS14 | 0.88 |
| 2 | 5 | rs4869314 | ERAP2 | ERAP2 | 0.82 |
| 3 | 1 | rs6679677 | RSBN1 | HIPK1 | 0.69 |
| 4 | 3 | rs34173901 | GLB1 | CCR4 | 0.65 |
| 5 | 3 | rs79815064 | CCR3 | CCRL2 | 0.64 |
| 6 | 3 | rs79815064 | CCR3 | CCR2 | 0.57 |
| 7 | 3 | rs79815064 | CCR3 | CCR3 | 0.51 |

### Functional annotation and CRISPRa/i validation of prioritised JIA SNPs

Risk loci often contain multiple variants in LD, making it essential to determine variant functionality to prioritise causal candidates. To achieve this, we overlapped all JIA-associated variants with ATAC-seq data from peripheral blood CD4+ T cells of three JIA patients to identify SNPs within accessible chromatin regions. To further assess their regulatory potential, these SNPs were annotated with EpiMap-predicted active chromatin regions (listed in Supplementary Table 2) across multiple immune cell types (listed in Supplementary Table 3). Additionally, gene expression correlations with these enhancer regions, as reported by EpiMap [18], were examined alongside RegulomeDB [31] functional scores (summarised in Supplementary Table 4 and 5). Overall, 614 out of 735 (84%) credible set SNPs and 238 out of 384 (62%) non-fine-mapped SNPs (LD R² > 0.8 to the lead SNP) overlapped EpiMap active enhancers in CD4+ T cells.

For further functional validation, we focused on loci containing COGS-prioritised genes. We selected loci with clear candidate SNPs, defined as those overlapping ATAC-seq peaks with a definitive target gene and numbering fewer than 10 candidate SNPs. Based on these criteria, we selected *CCR3*, *CCR4*, and *HIPK1* for functional studies, **(Table 3)**. In contrast, *RGS14* and *ERAP2* were excluded as they did not meet these prioritisation criteria, both loci contained numerous candidate SNPs, **shown in Supplementary Figure 1**.

**Table 3.**
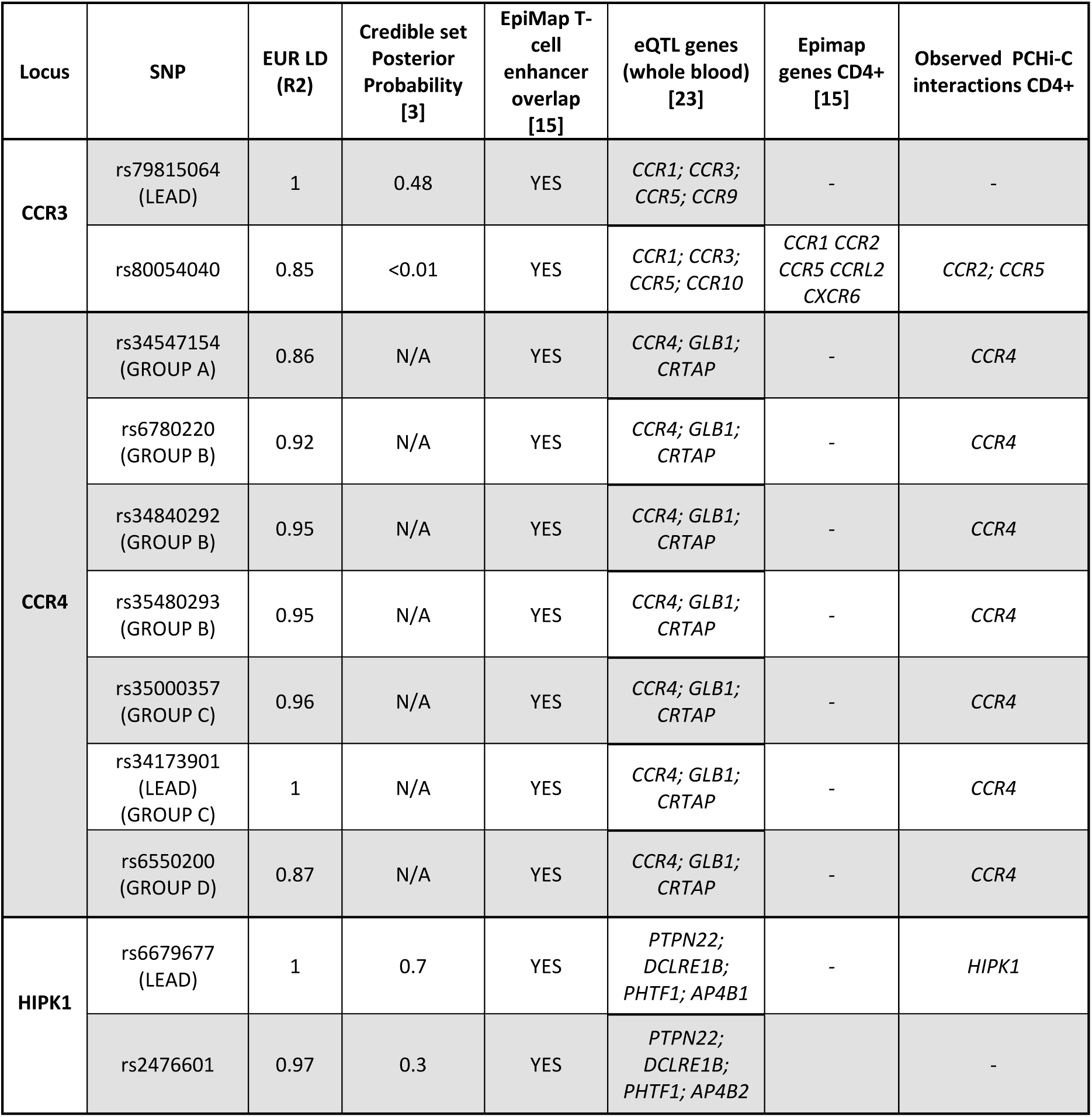
Summary of the genetic and functional annotations of JIA-associated SNPs within the *CCR3*, *CCR4*, and *HIPK1* loci.

#### CCR3

The CCR3 locus, shown in **Figure 1A**, contained three prioritised genes: *CCR2*, *CCR3*, and *CCRL2*. Out of 39 credible SNPs, 31 overlapped predicted T cell enhancers, with four also correlating with expression of surrounding genes in CD4+ T cells, as per EpiMap data (Supplementary **Table 4**). Among these, only rs80054040 overlapped an ATAC-seq peak in CD4+ T cells from JIA patients and demonstrated interactions with *CCR2* and *CCR5* (**Table** 3). Additionally, we followed up on rs79815064, which exhibited the highest posterior probability (0.48) from the credible SNP set.

**Figure 1.**
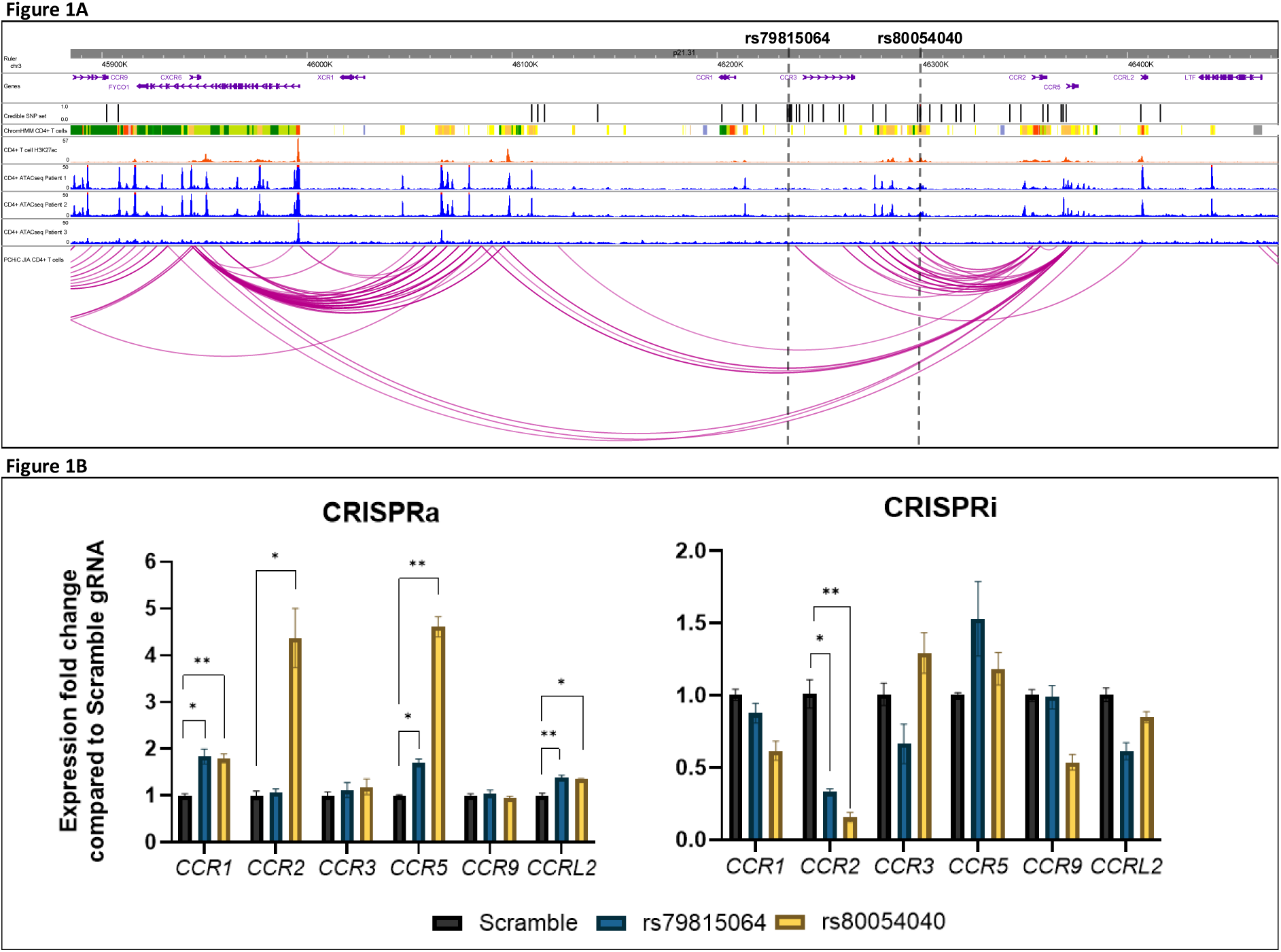
**A)** Prioritised risk locus *CCR3* overview. Tracks top to bottom 1) Ruler, 2) Genes (MANE Project), 3) JIA Credible SNPs, 4) chromHMM predicted chromatin state from peripheral blood T cells (E034), colour coded, described in Supplementary Table 3, 5) H3K27ac ChIP-seq T cells (Roadmap), 6-8) ATAC-seq CD4+ T cells JIA patient 1-3, 9) PCHi-C CD4+ T-cells JIA patients (3 patients merged) **B)** qPCR results of CRISPRa and CRISPRi in Jurkats targeting rs79815064 and rs80054040. Graphs show the mean fold change (±SEM) of cells containing SNP-targeting gRNAs compared to cells containing the scramble gRNA. Significance on the graph’s id indicated with asterisks: *: P<0.05; **: P<0.01; ***: P<0.001; ****: P<0.0001, no asterisk means no significant change was detected.

CRISPRa targeting of rs79815064 led to upregulation of *CCR1*, *CCR5*, and *CCRL2*. Similarly, targeting rs80054040 resulted in the same gene upregulation, with additional activation of *CCR2*. The strongest induction was observed for *CCR2* and *CCR5* when targeting rs80054040. Conversely, CRISPRi experiments at both SNP sites significantly reduced *CCR2* expression (**Figure** 1B). The most notable finding here is the robust regulatory effect on *CCR2* expression, as both SNPs were capable of upregulating and downregulating gene expression, strongly supporting their role as functional enhancer elements in T cell regulatory activity. *CCR2* and the other surrounding chemokine receptor genes that have previously been implicated in autoimmune disease. Elevated levels of the CCR2 ligand CCL2 has been observed in JIA patient’s synovial fluid, suggesting an active role for the CCR2– CCL2 axis in disease progression [32]. Therapeutically, *CCR2* and chemokine receptors in general have been a target of interest, with small-molecule antagonists and monoclonal antibodies designed to block its function provide potential drug targets [33].

#### CCR4

COGS also prioritised ***CCR4*** (COGS score = 0.65) (**Figure 2A**). The *CCR4* locus was not fine-mapped in the original GWAS study by López-Isac *et al*., [2]. All SNPs in the region mapped to introns of the Galactosidase beta 1 gene (*GLB1*) and overlapped predicted immune specific enhancers. No single candidate SNP emerged; thus, we chose to target all 7 SNPs in the region. The SNPs were divided into 4 groups (**Table 3**) based on proximity making them suitable for CRISPRa/i targeting.

**Figure 2.**
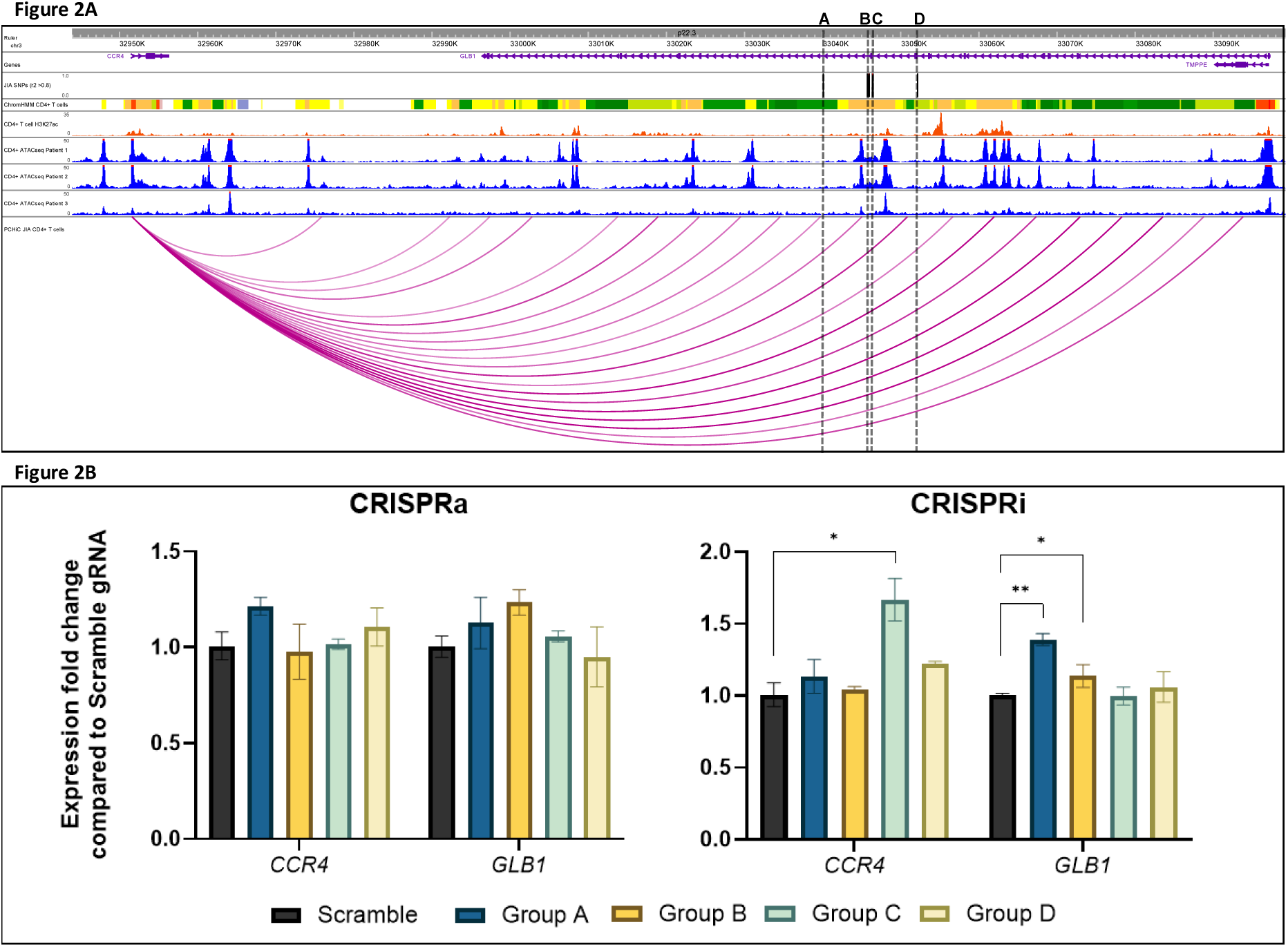
**A)** Prioritised risk locus *CCR4* overview. Tracks top to bottom 1) Ruler, 2) Genes (MANE Project), 3) JIA Credible SNPs, 4) chromHMM predicted chromatin state from peripheral blood T cells (E034), colour coded, described in Supplementary Table 3, 5) H3K27ac ChIP-seq T cells (Roadmap), 6-8) ATAC-seq CD4+ T cells JIA patient 1-3, 9) PCHi-C CD4+ T-cells JIA patients (3 patients merged) **B)** qPCR results of CRISPRa and CRISPRi in Jurkats targeting 4 SNP groups, listed in Table 3. Graphs show the mean fold change (±SEM) of cells containing SNP-targeting gRNAs compared to cells containing the scramble gRNA. Significance on the graph’s id indicated with asterisks: *: P<0.05; **: P<0.01; ***: P<0.001; ****: P<0.0001, no asterisk means no significant change was detected.

At the ***CCR4*** locus none of the CRISPRa experiments showed a difference in expression of either *CCR4* or *GLB1* regardless of SNP group targeted. On the other hand, CRISPRi samples targeting SNP group C showed some upregulation of *CCR4* expression. Additionally, targeting SNP groups A and D demonstrated an increase in expression of *GLB1* (**Figure 2B),** supporting a broader regulatory role for this region. There is a clear interaction between JIA SNPS and the *CCR4* gene promoter, however the modulation of *CCR4* here is not consistent to what was expected. This may indicate a complex feedback regulation or cell-specific effects that warrant further investigation. There is no current evidence to suggest a role for *GLB1* in autoimmunity whereas *CCR4* is a well-established autoimmune-associated gene, with a known role in immune cell trafficking and has been suggested to have a role in early disease based accumulation of *CCR4* + T cells in JIA synovial fluid [25].

#### HIPK1

In the ***HIPK1*** locus the lead SNP, rs6679677, is located between the round spermatid basic protein 1 (*RSBN1*) and the putative homeodomain transcription factor 1 (*PHTF1*) genes, shown in **Figure 3A**. It is in LD with just one other SNP, rs2476601, which resides within a protein tyrosine phosphatase non-receptor 22 (*PTPN22*) exon, causing a non-synonymous change. The intergenic variant rs6679677, had the higher posterior probability in the credible SNP set of 0.7 and overlaps an EpiMap predicted CD4+ T cell weak enhancer and does not overlap any ATAC-seq peaks in CD4+ T cells. rs2476601 is well studied and has already been shown to be involved in immunity through *PTPN22* [34,35]. Here we were interested in understanding if rs667977 could also have a regulatory role on additional putative candidate genes in this region, given it had a higher posterior probability and if it could affect COGS prioritised gene, *HIPK1*.

**Figure 3.**
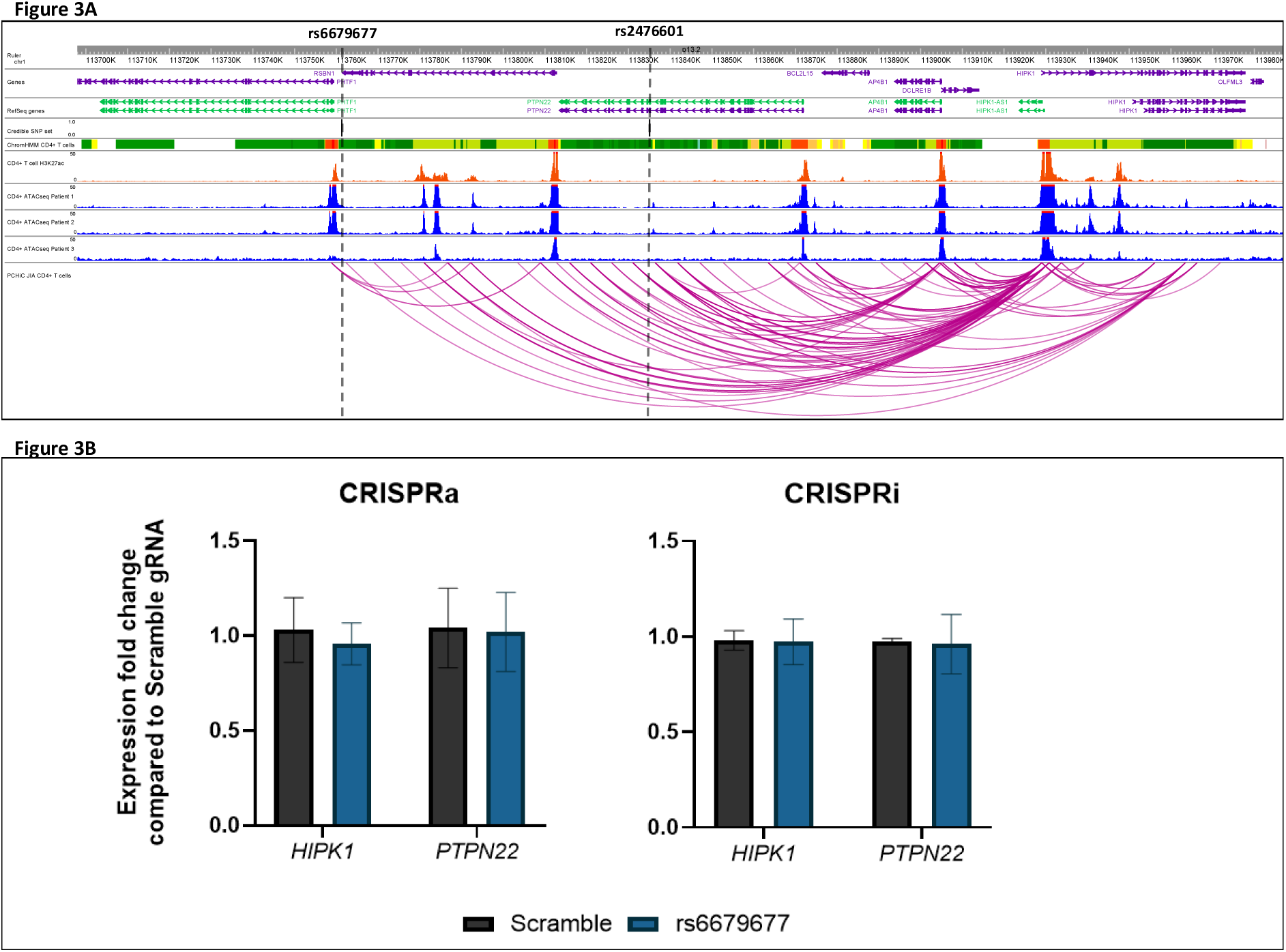
**A)** Prioritised risk locus *HIPK1* overview. Tracks top to bottom 1) Ruler, 2) Genes (MANE Project), 3) JIA Credible SNPs, 4) chromHMM predicted chromatin state from peripheral blood T cells (E034), colour coded, described in Supplementary Table 3, 5) H3K27ac ChIP-seq T cells (Roadmap), 6-8) ATAC-seq CD4+ T cells JIA patient 1-3, 9) PCHi-C CD4+ T-cells JIA patients (3 patients merged) **B)** qPCR results of CRISPRa and CRISPRi in Jurkats targeting rs6679677. Graphs show the mean fold change (±SEM) of cells containing SNP-targeting gRNAs compared to cells containing the scramble gRNA. Significance on the graph’s id indicated with asterisks: *: P<0.05; **: P<0.01; ***: P<0.001; ****: P<0.0001, no asterisk means no significant change was detected.

At the ***HIPK1*** locus, targeting of rs6679677 in both CRIPSRa/i Jurkat cell lines did not result in any modulation of either *HIPK1* or *PTPN22* **(Figure 3B)**. These findings suggest that rs6679677 does not play a significant regulatory role in *HIPK1* or *PTPN22* expression, reinforcing the likelihood that rs2476601, a well-characterised *PTPN22* coding variant, is the primary causal SNP at this locus.

## Discussion

Despite efforts to understand JIA biology, the precise mechanisms underlying the disease remain only partially understood. Recent GWAS have provided valuable insights in understanding JIA. However, GWAS studies leverage LD to identify genetic risk regions, and risk loci often contain numerous SNPs in LD, many of which have no functional impact. Distinguishing true causal variants remains a key hurdle. Additionally, these risk SNPs primarily influence regulatory functions rather than protein-coding sequences, further complicating target gene identification. Previous JIA research has utilised transcriptomics-based approaches to identify disease-associated genes. Studies such as Zhang *et al*., and Feng *et al*., used TWAS to infer causal genes based on genetically regulated expression across tissues, and has led to identifying hundreds of differentially expressed genes [36,37]. Other studies like Tarbell *et al*., and Jiang *et al*., incorporated epigenetic factors and public 3D chromatin interaction data to prioritise GWAS SNPs and identify candidate genes [38,39]. Here, we present the first gene-promoter interaction map in CD4+ T cells from JIA patients, leveraging a low-input application of PCHi-C. We integrated COGS to prioritise regulatory interactions between JIA risk SNPs and their target genes in CD4+ T cells. This identified additional candidate genes beyond those highlighted in the López-Isac *et al*., GWAS. Using CRISPRa/i for functional validation, we identified novel SNP-gene relationships and provided supporting evidence for candidate causal genes.

In the *CCR3* locus, two SNPs, rs80054040 and rs79815064, were prioritised for their regulatory potential. While rs79815064 had the highest posterior probability score from the credible set, it showed mild influence on expression of surrounding genes underscoring the importance of empirical validation. rs80054040 demonstrated a clear regulatory role, particularly in the upregulation of *CCR2* and *CCR5* when targeted with CRISPRa. CCR2 and CCR5 are chemokine receptors found primarily on the cell surface of leukocytes. They’re involved in the control of leukocyte migration, largely monocytes, macrophages and T cells, through detection of chemokine gradients [40]. This chemokine receptor rich locus has been previously implicated in the pathogenesis of several autoimmune diseases such as multiple sclerosis, rheumatoid arthritis and systemic lupus erythematosus, underscoring their pivotal role in immune regulation [41–43].

The significance of these genes in various diseases is underscored by the development of drugs that specifically target them [44–46]. CCR2 and CCR5 are considered promising therapeutic targets due to their critical roles in mediating the migration and recruitment of immune cells to inflamed joints. A widely used RA treatment, Methotrexate, doesn’t directly bind these receptors, but leads to the downregulation of *CCR2* expression on monocytes and CD4+ T cells in RA patients [47]. It was hypothesised that by targeting these receptors directly, it may be possible to reduce synovial inflammation and joint destruction. Antagonists targeting CCR2 and CCR5 have been employed in animal models that mimic RA-like conditions, exhibiting arthritic characteristics and an increased expression of chemokine ligands and receptors have shown positive results with reduced joint inflammation [48–50]. A clinical trial of an anti-CCR2 blocking antibody in RA patients led to a decrease of CCR2 on monocytes by at least 56%; however, it did not reduce inflammation in the joints [7]. Similarly, CCR5 antagonists in RA also failed to demonstrate alleviation of any RA related symptoms [6,51]. The failure to observe a positive outcome from the clinical trials of CCR2 and CCR5 antagonists may be attributed to the redundancy of the chemokine system. Since there are multiple receptors that can facilitate the migration of immune cells into the synovium, blocking CCR2 and CCR5 alone might not be sufficient. This complexity in the chemokine system may explain why targeting these specific receptors did not yield the desired therapeutic results in RA. It’s important to highlight that the clinical trials mentioned were conducted in the context of RA, not JIA. Intriguingly, the most recent multi ancestry GWAS in RA did not identify any genetic links to this locus [52], unlike in JIA [2]. This discrepancy suggests that the roles of these receptors likely differ between the two diseases.

Investigating the function and impact of CCR2 and CCR5 in the context of JIA, which has a known genetic association with this locus, could provide valuable insights and may be a worthwhile avenue for future research.

At the *HIPK1* locus, candidate SNP rs6679677, did not exhibit a discernible regulatory role, suggesting that the exonic SNP in LD, rs2476601 affecting *PTPN22* is the likely causal variant. This aligns with existing literature implicating *PTPN22* in autoimmune pathogenesis [35,53]. While a recent study, identified *HIPK1* as a potential causal gene in JIA, through interactions with rs6679677 and showing *HIPK1* downregulated in JIA patients, this locus requires further functional characterisation to identify the role of *HIPK1* in JIA [28]. At the *CCR4* locus, the only locus here of suggestive genome-wide significance, all 7 risk SNPs were targeted. CRISPRa did not reveal any meaningful gene modulations whereas targeting with CRISPRi led to significant increases of *CCR4* and *GLB1* in certain instances, this must be interpreted with caution given the relative low baseline expression of *CCR4* and *GLB1* in Jurkats which may inflate significance, given that observed fold changes ranged from 1.21 to 1.66. This may also point to a complex regulatory mechanism in the region.

While this study advances our understanding of JIA pathogenesis and the genetic regulation, several limitations remain. The analysis of CD4+ T cells does not account for the heterogeneity among specific subpopulations, such as Tregs, Th1, and Th17 cells, which have distinct roles in autoimmunity and could provide deeper insights if analysed separately [54,55]. Additionally, the focus on peripheral blood-derived CD4+ T cells may not fully capture the unique gene expression profiles and chromatin dynamics of T cells within the synovial tissue, the primary site of inflammation in JIA [56].

In conclusion, this study underscores the complexity of linking GWAS-identified SNPs to their functional roles and target genes in JIA. The integration of genomic and epigenomic data can facilitate the prioritisation of candidate risk SNPs, enabling the validation of their functional roles and the identification of potential risk genes and novel therapeutic targets. Using a low-input PCHi-C approach combined with COGS, we identified, *CCR2*, *CCR3*, *CCRL2*, *ERAP2*, *RGS14*, *HIPK1* and *CCR4* as putative causal genes in JIA. Furthermore, through CRISPRa/i validation, we provide strong evidence that rs80054040 and rs79815064 function as regulatory elements modulating *CCR2* and *CCR5* expression, both of which hold therapeutic potential in JIA.

## Acknowledgments

This study acknowledges the use of the UK JIA cohort Childhood Arthritis Prospective Study (CAPS) (funded by Versus Arthritis, grant reference number 20542). We express our gratitude to the patients, nurses, study coordinators, and investigators who were involved in CAPS. The authors would also like to acknowledge the assistance given by IT Services and the use of the Computational Shared Facility at The University of Manchester.

## Contributors

Study conception: AF, GO. Contribution to the overall study design: AF, VM, SE, WT, JB, MS, GO. Acquisition of data: AF, VM, DZ, MG, SR, JD. Performed the statistical analyses: VM, AF, CS. Writing the first draft of the manuscript: AF, VM. Funding acquisition: GO, SE, WT, JB, MS. All authors critically read, reviewed and approved the final manuscript.

## Data availability statement

Data are available on reasonable request.

## Patient and public involvement

Patients and/or the public were not involved in the design, or conduct, or reporting, or dissemination plans of this research.

